# Economic Analysis of 15-valent and 20-valent Pneumococcal Conjugate Vaccines among Older Adults in Ontario, Canada

**DOI:** 10.1101/2024.10.04.24314906

**Authors:** Gebremedhin B. Gebretekle, Ryan O’Reilly, Stephen Mac, Shaza Fadel, Natasha S. Crowcroft, Beate Sander

## Abstract

**Introduction:** Despite the availability of publicly funded 23-valent polysaccharide pneumococcal vaccine (PPV23) for all adults aged 65 years and older, pneumococcal disease remains a public health concern in Ontario, Canada. Health Canada approved two new pneumococcal conjugate vaccines (PCV), 15-valent (PCV15) and 20-valent (PCV20) and we assessed their cost-effectiveness for older Ontarians.

**Methods:** We conducted a cost-utility analysis using an individual-level state transition model to compare one dose of PCV (alone or in series with PPV23) with PPV23-only. We estimated sequential incremental cost-effectiveness ratio (ICER) expressed in costs (C$2022) per quality-adjusted life years (QALYs) from the healthcare payer perspective, discounted at 1.5% per annum. We performed sensitivity and scenario analyses to examine impact parameter uncertainties on the results.

**Results:** A sequential comparison of vaccination strategies with no indirect effect from childhood vaccination resulted in an ICER of $44,324/QALY for PCV15-alone compared to PPV23-only, and $70,751/QALY for PCV20-alone versus PCV15-alone. None of the PCV15/20 combined with PPV23 programs were cost-effective at a cost-effectiveness threshold of C$50,000/QALY. PCV20 alone had an ICER of C$46,961/QALY compared to PPV23-only. When considering the indirect effects, use of PCV15/20 alone or in series with PPV23 were not cost-effective strategies. ICERs were mostly influenced by vaccine characteristics (effectiveness, waning, cost) and the incidence of pneumococcal community-acquired pneumonia.

**Conclusion:** Vaccinating older adults with PCV15/20 is likely to reduce burden of pneumococcal disease and would be cost-effective initially, but is expected to be less economically attractive in the longer-term when herd immunity benefits from childhood vaccination programs are considered.

## INTRODUCTION

*Streptococcus pneumoniae* (pneumococcus) is a gram-positive bacterium with 100 known serotypes that causes a wide spectrum of disease, manifesting either as invasive pneumococcal disease (IPD; e.g., meningitis, bacteremia) or noninvasive disease (e.g., non-bacteremic pneumococcal pneumonia (NBP)) [1-3]. The pathogen remains the leading cause of morbidity and mortality worldwide including Canada, particularly affecting young children and adults aged ≥ 65 years (hereinafter referred to as “older adults”) [4, 5]. In Ontario, 42% of IPD cases in 2019 were among older adults and 87% of these cases were hospitalized [4].

To reduce the burden of disease, Ontario has long implemented age- and risk-based pneumococcal vaccination programs [6]. The 23-valent pneumococcal polysaccharide vaccine (PPV23) has been publicly funded for all older adults (regardless of risk factors) for decades, whereas PCV13 is only publicly funded for adults with immunocompromising conditions [6]. Despite these vaccination efforts, the burden of pneumococcal diseases among these population remains high. From 2007 to 2018, the incidence rate of PPV23/non-PCV13 serotypes increased from 2.6 to 3.9 per 100,000 people, and non-vaccine serotypes rose from 1.5 to 2.4 per 100,000 people [7, 8]. The health and economic burden of pneumococcal disease could further increase in the future, not only because of the progressively ageing population but also due to the rapid spread of resistant *S. pneumoniae* strains. Between 2014 and 2018, the proportion of vaccine-preventable (PCV13) multidrug-resistant (MDR) invasive *S. pneumoniae* infections in Canada increased by 25% (from 9.2% to 11.5%), while MDR isolates identified in non-vaccine preventable (non- PCV13) IPD cases increased by 74% [9].

Although studies have shown that PPV23 is effective in preventing IPD, its effectiveness against non-invasive (non-bacteremic) pneumococcal disease remains uncertain [10-13]. Pneumococcal polysaccharide conjugate vaccines (in which a bacterial polysaccharide is covalently conjugated to an immunogenic carrier protein) are effective in preventing both IPD and pneumococcal pneumonia [14-16]. Hence, vaccination of older adults with higher-valency pneumococcal conjugate vaccines (PCV) appears to be the most promising public health intervention for prevention of pneumococcal disease and its associated burden. Health Canada has approved two new PCV for adults: the 15-valent pneumococcal conjugate vaccine (PCV15) and the 20-valent pneumococcal conjugate vaccine (PCV20) [17]. PCV15 contains all serotypes included in PCV13 (1, 3, 4, 5, 6A, 6B, 7F, 9V, 14, 18C, 19A, 19F, and 23F), plus two additional serotypes (22F and 33F), with the non-PCV13 serotypes accounting about 15% of IPD in older adults [18]. Likewise, PCV20 adds seven non-PCV13 serotypes (8, 10A, 11A, 12F, 15B, 22F and 33F), which account for 41.7% of IPD in this population [18]. These vaccines could help to address important public health needs by providing broader coverage for serotypes associated with a substantial proportion of pneumococcal disease. The National Advisory Committee on Immunization (NACI) in Canada has recommended a single dose of PCV20 for all older adults, with the option of using PCV15 plus PPV23 if PCV20 is unavailable [19] and Ontario Immunization Advisory Committee has adopted this recommendation [6]. We conducted a model-based economic evaluation to assess the cost-effectiveness of various pneumococcal vaccination strategies for older adults in Ontario.

## METHODS

We conducted a model-based cost-utility analysis to assess the cost-effectiveness of five potential pneumococcal vaccination strategies for older adults in Ontario, Canada: (i) PCV15-alone, (ii) PCV20- alone, (iii) PCV15 in series with PPV23, (iv) PCV20 in series with PPV23, and (v) PPV23-only (current standard of care). The recommended interval between PCV15/20 and PPV23 vaccination is at least one year, assuming that conjugated vaccine would be given first [19]. An individual-level microsimulation model was used to predict the lifetime costs and health outcomes of these interventions from the healthcare payer perspective. Health outcomes included the expected cumulative number of individuals with IPD and community-acquired pneumonia (CAP) (outpatient and inpatient), number of deaths due to pneumococcal disease, and quality adjusted life years (QALYs). QALYs gained were determined by multiplying the time (in years) lived in a given health state with corresponding utility weights [20]. We computed a sequential incremental cost-effective ratio (ICER), defined as incremental cost per incremental health effects (C$/QALY gained) for all strategies. QALYs and costs were discounted at 1.5% per annum [21, 22].

### Model Structure

Based on a previous model developed by our team [23], we expanded and updated the microsimulation model with a 1-year cycle length (i.e., time step) comprising three mutually exclusive health states: healthy, sequelae due to *S. pneumoniae* infection, and death, and acute events occurring prior to transition from “healthy” to “sequelae” or “death” (Figure 1). Individuals enter the model in the “healthy” state at the age of 65 years and, over time, they could be infected with *S. pneumoniae* and could develop pneumococcal disease (IPD-meningitis, IPD-bacteremia, outpatient or inpatient NBP), or remain in the “healthy” state. We assumed that all patients with IPD would be hospitalized or seek emergency care as it is an acute and serious disease in older adults. Individuals who experienced pneumococcal disease could recover and return to the “healthy” state, or progress to post-meningitis “sequelae”, or “death” state. We assumed that individuals who were diagnosed with outpatient CAP could only die from causes other than pneumococcal infection. Only individuals with IPD-meningitis could develop permanent disability or sequelae, manifested as neurologic impairment or hearing loss. We assumed individuals in the “sequelae” state would be at the same risk of contracting subsequent pneumococcal infections as individuals in the “healthy” state. Individuals who transitioned to sequelae could die or remain impaired. We assumed that individuals can only have one sequelae episode over their lifetime, given that the occurrence of more than one sequelae is very rare (less than one per million [24]). Individuals could die from causes other than pneumococcal infection at any time. Vaccination modified the risk of *S. pneumococcal* infection but did not affect disease severity once infected. We did not consider revaccination assuming that individuals would be vaccinated either a single vaccine or a complete series once in their lifetime. We developed the model using TreeAge Software (TreeAge Software, Inc., Williamstown, MA).

**Figure 1.**
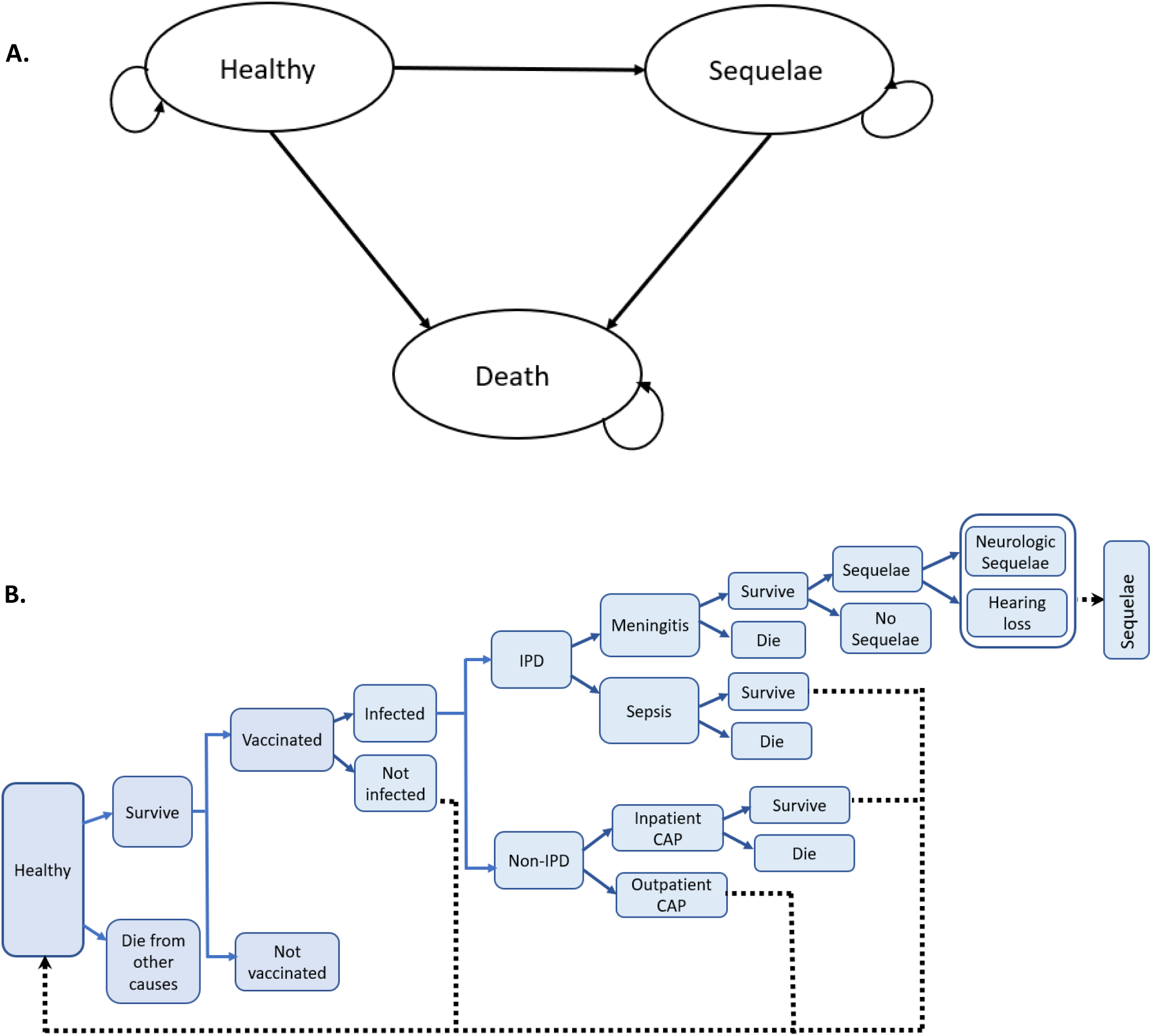
Model schematic. **A**. Markov state transition diagram depicting the natural history of pneumococcal infection and disease among immunocompetent older adults. The circles show possible health states while the arrows imply transitions of individuals among different health states. During each cycle, individuals could be in any of the health states and the transitional probabilities determine their possible transition between health states. **B**. Simplified model structure for acute events in the “healthy” state. CAP: Community-Acquired Pneumonia; IPD: Invasive pneumococcal disease.

### Parameter Values

Model parameter values, including disease incidence, mortality, vaccine effectiveness, utilities, and costs are summarized in Table 1. Most parameter values were informed by Ontario health administrative and surveillance data, supplemented by published literature.

**Table 1.** Model parameter values.

| Parameter | 65-74 years | 75-84 years | 85+ years | Source |
| --- | --- | --- | --- | --- |
| Annual Disease Incidence (per 100,000 population) |  |  |  |  |
| IPD - Meningitis | 0.8 | 1.1 | 2.1 | iPHIS, 2017-2019 [23] |
| IPD - Bacteremia | 14.5 | 19.5 | 39.2 | iPHIS, 2017-2019 [23] |
| Inpatient CAP | 282 | 745 | 2,027 | ICES, 2017-2019 [23] |
| Outpatient CAP | 2,081 | 3,097 | 4,981 | ICES, 2017-2019 [23] |
| Proportion of Sequelae after meningitis (%) | 31.7 | 31.7 | 31.7 | Jit,2010 [26] |
| Annual Case Fatality Rate (%) |  |  |  |  |
| IPD | 19 | 29 | 46 | ICES, 2017-2019 [23] |
| Inpatient CAP | 12 | 17 | 24 | ICES, 2017-2019 [23] |
| Background Mortality Rate | Age-Specific | Age-Specific | Age-Specific | Statistics Canada [28] |
| Vaccine Effectiveness (%) |  |  |  |  |
| PCV against IPD | 0.89 | 0.76 | 0.44 | Van Werkhoven *et al*,2015 [15] |
| PCV against ST3-IPD | 26 | 26 | 26 | Stoecker,2021 [44] |
| PCV against NBP | 0.60 | 0.37 | 0.20 | Van Werkhoven *et al*,2015 [15] |
| PPV23 against IPD | 73 | 73 | 73 | Falkenhorst *et al*,20117 [29] |
| PPV23 against ST3-IPD | 2 | 2 | 2 | Djennad *et al*, 2018 [45] |
| PPV23 against NBP | 0.0 | 0.0 | 0.0 | Moberley *et al*,2013 [13] |
| Costs (C\$) | | | | |
| IPD - Meningitis | 26,961 | 22,584 | 9,832 | ICES, 2017-2019 [23] |
| IPD - Bacteremia | 26,961 | 22,584 | 22,584 | ICES, 2017-2019 [23] |
| Inpatient CAP | 20,297 | 17,985 | 11,977 | ICES, 2017-2019 [23] |
| Outpatient CAP | 781 | 1,176 | 1,115 | ICES, 2017-2019 [23] |
| Healthy (baseline costs) | 4,733 | 7,770 | 13,106 | ICES, 2017-2019 [23] |
| Sequelae | 1,561 | 1,561 | 1,561 | Assumption |
| Vaccine |  |  |  |  |
| PPV23 Price per dose | 11 | 11 | 11 | Atwood *et al*, 2018 [37] |
| PCV15 Price per dose | 97 | 97 | 97 | CDC,2022 [38] and Private Pharmacy price (personal communication) |
| PCV20 Price per dose | 112 | 112 | 112 |  |
| Vaccine Administration Cost (1st dose) | 5.40 | 5.40 | 5.40 | OHIP Schedule of Benefits [39] |
| Vaccine Administration cost (2<sup>nd</sup> dose) | 38.00 | 38.00 | 38.00 | OHIP Schedule of Benefits [39] |
| Utilities |  |  |  |  |
| IPD - Decrement | 0.0709 | 0.0709 | 0.0709 | Assumption |
| Inpatient CAP - Decrement | 0.0709 | 0.0709 | 0.0709 | Mangen *et al*,2017 [36] |
| Outpatient CAP - Decrement | 0.0062 | 0.0062 | 0.0062 | Oppong *et al*,2018 [35] |
| Sequelae - Decrement | 0.26 | 0.26 | 0.26 | Oostenbrink *et al*,2002 [46] |
| Healthy | Age-Specific | Age-Specific | Age-Specific | Guertin *et al*,2018 [34] |

CAP= Community-acquired pneumonia; CDC=Centers for Disease Control and Prevention; IPD=Invasive pneumococcal disease; iPHIS = Integrated Public Health Information System; PCV15 = 15-valent pneumococcal conjugate vaccine; PCV20 = 20-valent pneumococcal conjugate vaccine; PPV23 = 23-valent pneumococcal polysaccharide vaccine

### Pneumococcal Disease Incidence and Mortality

The number of IPD cases by serotype classifications and age group were obtained from Ontario’s integrated Public Health Information System (iPHIS) database for the years 2007 to 2019 and used to calculate annual IPD incidence rates by age group (65-74, 75-84, 85+ years). IPD cases by vaccine-specific serotypes were grouped into the following categories: serotypes covered by PCV13 (1, 3, 4, 5, 6A, 6B, 7F, 9V, 14, 18C, 19A, 19F, and 23F), PCV15/non-PCV13 (22F and 33F), PCV20/non-PCV15 (8, 10A, 11A, 12F, and 15B), PPV23/non-PCV20 (2,9N, 17F,20) and non-vaccine types (all serotypes not included in PCV13, PCV15, PCV20 and PPV23). Population data from Statistics Canada was used to convert episode counts to annual incidence rates [25]. IPD incidence rates increased with age, and the proportion of IPD- bacteremia was higher than IPD-meningitis in all age groups (Table 1). The proportion of sequela following pneumococcal meningitis (31.7%) was derived from published Ontario data [26]. All-cause CAP incidence rates was derived using Ontario’s population-level health administrative database held at ICES [23]; and then calculated the incidence of CAP due to *S. pneumoniae* based on a Canadian study that showed 10.1% of all-cause CAP is attributable to *S. pneumoniae*, with ratios being constant over time [27]. Age-specific case-fatality rates for inpatient CAP were derived from the same database. We assumed that outpatient CAP does not result in death. IPD attributed deaths were also estimated based on ICES data [23]. We assumed that overall incidence for all serotypes would remain stable throughout the time horizon. Age-specific all-cause mortality rates were obtained from Statistics Canada [28].

### Vaccine Effectiveness

The effectiveness of PPV23 against IPD in older adults was estimated to be 73% (60-80%), based on a meta-analysis of published studies [29]. Consistent with other published studies, our base-case analysis assumed that PPV23 is ineffective in preventing all-cause NBP [12, 13]. Effectiveness of PPV23 wanes over time, assuming a linear decline and reaching zero five years post-vaccination [30]. In the absence of effectiveness data for PCV15/20, we assumed non-inferior immunogenicity of the PCV13 conjugate vaccine. The effectiveness of PCV13 in preventing vaccine-type IPD and NBP was based on the *post-hoc* analysis of the Community-Acquired Pneumonia immunization Trial in Adults (CAPiTA) [15]. We projected that both PCV15 and PCV20 would provide protection for 15 years, with full effectiveness for the first 5 years, followed by a linear decline to zero over the subsequent 10 years [16]. The base case analysis did not include potential indirect effects of routine childhood vaccination programs due to limited data availability. A scenario analysis was conducted to account for the indirect effects, assuming a 50% linear reduction in pneumococcal disease caused by unique serotypes in PCV15/20 over five-year period. This assumption is based on previous studies on the PCV13 vaccine and is consistent with the approach used in the model-based economic evaluation by the Public Health Agency of Canada [31-33].

### Health State Utilities

Age-specific utilities for individuals who did not develop pneumococcal diseases were derived from the Canadian National Population Health Survey measured using Health Utilities Index II [34]. The utility weight for outpatient CAP was obtained from European outpatients with lower respiratory tract infections measured using the EuroQol five-dimensional questionnaire (EQ-5D) [35]. Utility weights for patients with inpatien*t* CAP were derived from a case-control study nested within CAPiTA, using EQ-5D [36]; and we assumed the same utility weight for inpatients with IPD.

### Costs

Costs included vaccination program costs, and inpatient and outpatient care costs for pneumococcal disease from the healthcare payer perspective. Vaccine prices for public payers in Canada are confidential. We estimated the price of PPV23 at C$11 per dose, assuming a 30% discount of price for private payers, to approximate the confidential government contract price [37]. The price of PCV15/20 was calculated based on the United States (US) Centers for Disease Control and Prevention pricing relative to PCV13 [38]. With PCV13 priced at C$126 per dose for private payers in Toronto, Ontario (University Health Network Pharmacy, personal communication), and applying a 30% discount, we estimated the per dose price of PCV15 at C$97 and PCV20 at C$112. Vaccine administration cost for the first dose was estimated at C$5.40, assuming it is given during a scheduled visit, while the second dose, administered after at least a one-year interval during a separate visit, was estimated at C$35 [39].

Patient-level medical costs, including physician services, ambulatory care, hospitalizations, diagnostics, medication, and care for chronic sequelae were estimated from Ontario’s health administrative data [23]. The study used a matched cohort study design to estimate population-based real-world attributable healthcare costs by disease episode. Thus, the average absolute cost of being in the unexposed group (baseline healthcare costs unrelated to pneumococcal disease, hereafter called “healthy cost”), as well as the cost of individuals with pneumococcal disease (i.e., costs of IPD, inpatient and outpatient CAP) were estimated. Costs of vaccine adverse events following immunization were not included, assuming that they are mild and unlikely to require healthcare services. All costs are in 2022 Canadian dollars and adjusted as needed using the Canadian Consumer Price Index [40].

### ANALYSIS

In the base case analysis, we considered a cohort of older adults living in Ontario being vaccinated at the age of 65 years. Future serotype distribution was assumed to be unaffected by potential indirect effects from children vaccinated with PCV15/PCV20. We ran 100,000 simulations to estimate the sequential ICERs by comparing all strategies against each other. Sequential ICERs were calculated by ranking all strategies by cost and excluding strategies that are strictly dominated (more costly and less effective) or subject to extended dominance (more costly and less effective than a combination of other strategies). ICERs for PCV15/20 alone compared to PPV23 were also calculated to allow for comparison with other studies that did not perform sequential analysis. In the absence of an established cost-effectiveness threshold in Canada, cost-effectiveness was assessed against a commonly used threshold of C$50,000/QALY.

One-way sensitivity analyses were performed to assess the impact of parameter uncertainties on the base case findings. In this analysis, values of each parameter such as vaccine cost, vaccine effectiveness, disease incidence, and treatment costs were changed one at a time over a plausible range of values while all other parameters were kept constant. Parameters were ranked according to ICER variation, and the most influential parameters are shown in a tornado diagram. Additionally, we explored the potential impact of indirect effects from pediatric vaccination programs through a scenario analysis.

## RESULTS

Vaccinating older adults with PCV15 or PCV20 was more effective at preventing pneumococcal disease-related outcomes than the current PPV23 vaccination program. Use of PCV15-alone resulted in a decrease of 36 IPD episodes (453 vs. 489), 355 CAP episodes (52,250 vs. 52,605), and 15 deaths (2,763 vs. 2,778) over the lifetime of 100,000 individuals when compared to PPV23-only. Likewise, use of PCV20-alone instead of PPV23 could prevent 46 IPD episodes (443 vs. 489), 389 CAP episodes (52,216 vs. 52,605), and 18 deaths (2,760 vs. 2,778).

Mean costs, QALYs and ICERs are presented in Table 2. Our base case analysis showed that vaccinating older adults with PCV15/20 (alone or in series with PPV23) resulted in more QALYs gained, but at a higher cost than PPV23-alone. When all strategies were compared sequentially, PCV15-alone was associated with an incremental expected cost of C$78 and incremental benefits of 0.001769 QALY per person compared to PPV23, resulting in an ICER of C$44,324/QALY. The ICER for PCV20-alone versus PCV15-alone was C$70,751/QALY. A combined use of PCV15 plus PPV23 was extendedly dominated by PCV20 plus PPV23, and therefore, excluded from the sequential ICER calculation. Using PCV20 in series with PPV23 instead of PCV20-alone resulted a higher ICER (C$958,988/QALY).

**Table 2.** Mean quality-adjusted life years gained, cost and incremental cost-effectiveness ratios, base case (with no indirect effects) and scenario analysis (with indirect effects).

| Strategy | Cost, C\$ | Effects (QALY) | ICER (C\$/QALY) | |
| --- | --- | --- | --- | --- |
|  |  |  | Sequential | Versus PPV23-only |
| Base-case analysis, no indirect effects |  |  |  |  |
| PPV23-alone | 118,152 | 15.216002 | - | - |
| PCV15-alone | 118,231 | 15.217771 | 44,324 | 44,324 |
| PCV20-alone | 118,245 | 15.217967 | 70,751 | 46,961 |
| PCV15 plus PPV23 | 118,276 | 15.217983 | Extended dominance | 62,464 |
| PCV20 plus PPV23 | 118,290 | 15.218015 | 958,988 | 68,632 |
| Scenario analysis, with indirect effects |  |  |  |  |
| PPV23-alone | 118,137 | 15.216675 | - | - |
| PCV15-alone | 118,219 | 15.217972 | Extended dominance | 63,223 |
| PCV20-alone | 118,234 | 15.218267 | 60,930 | 60,930 |
| PCV15 plusPPV23 | 118,264 | 15.218258 | Dominated | 84,227 |
| PCV20 plus PPV23 | 118,281 | 15.218289 | 2,136,364 | 89,219 |

We further compared PCV15 or PCV20 (alone or in series with PPV23) vaccination programs against a common baseline or denominator i.e., PPV23-only (Table 2, last column). The analysis showed that using PCV15 or PCV20 alone resulted ICERs of C$44,324/QALY and C$46,961/QALY, respectively. When PPV23 was added to PCV15 or PCV20, the ICERs increased to C$62,464/QALY and C$68,632/QALY, respectively.

When indirect effects from a childhood vaccination program were considered, PCV15 strategies were dominated or extendedly dominated by PCV20 strategies. Using PCV20-alone resulted an ICER of C$61,015/QALY gained compared to PPV23-only (Table 2). The combined use of PCV20 plus PPV23 generated higher ICER, both when compared to PCV20-alone (ICER of C$2,136,364/QALY) and PPV23- only (ICER of C$89,219/QALY).

The findings of the one-way sensitivity analyses are presented in Figures 2, 3 and 4. Compared to PPV23- alone, the use of PCV15 or PCV20 alone appears to be not cost-effective at a threshold of C$50,000/QALY when the PCV15 or PCV20 vaccine price increased to C$250 (vs. C$97 base cost of PCV15 or C$112 for PCV20), PCV15/20 had no effect against NBP (vs. 60%), the proportion of CAP attributed to *S. pneumoniae* decreased to 5% (vs. 10.1%), discount rate increased to 5% (vs. 1.5%), PPV23 effectiveness against NBP increased to 27.4% (vs. 0%), PCV15/20 effectiveness against IPD decreased to 40% (vs.89%) and treatment costs doubled. Compared to PCV15-alone, the use of PCV20- alone appears to be cost-effective at a threshold of <C$50,000/QALY when PCV20 vaccine price decreased to C$30 (vs. C$112 base cost) or PCV15 price increased to C$250 (vs. base case C$97) (Figure 4). Assuming PCV15 price of C$97, the threshold price at which PCV20 becomes cost-effective was calculated to be C$97.45.

**Figure 2.**
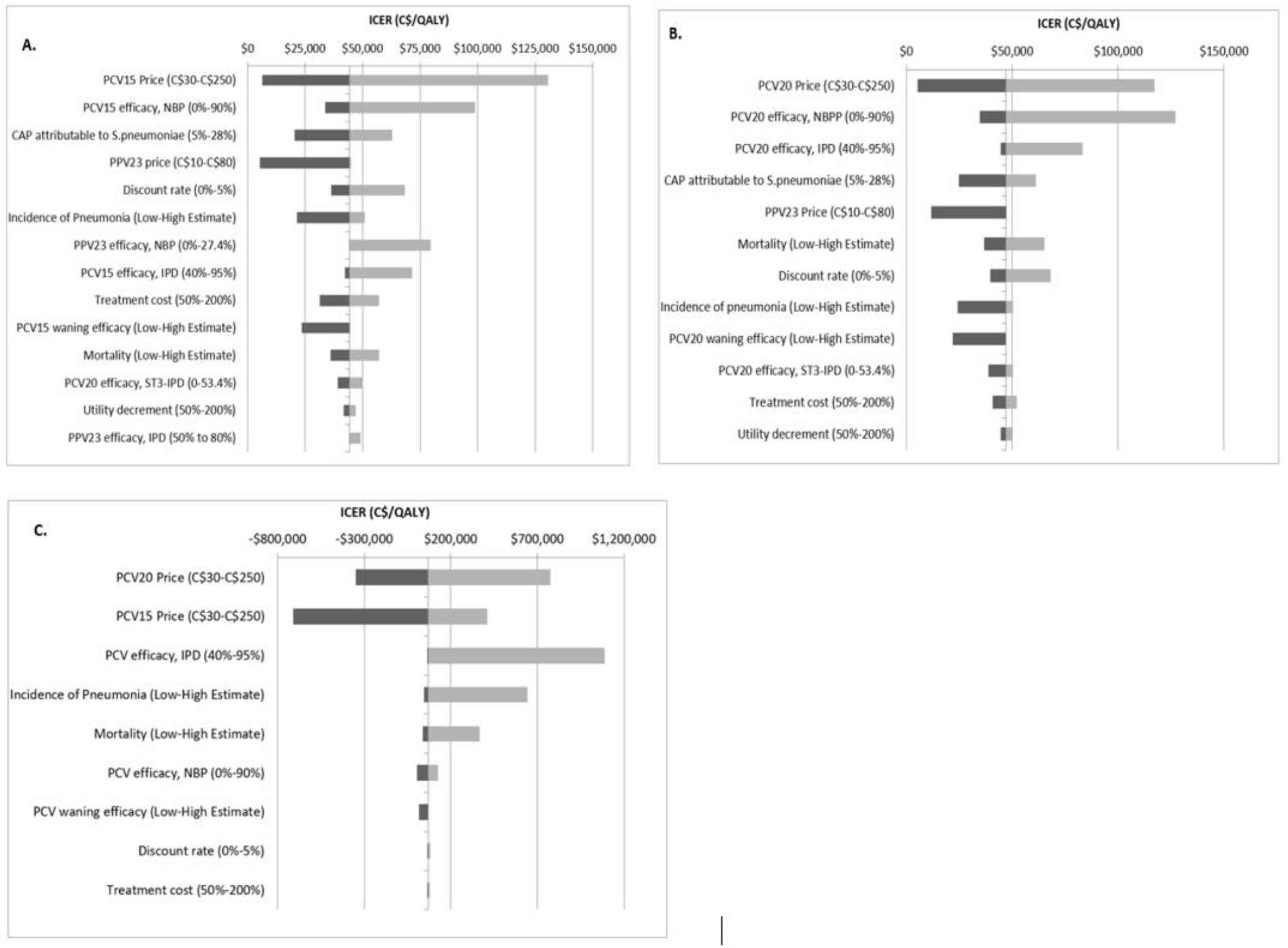
One-way sensitivity analyses result of key parameters comparing: **A**. PCV15-alone vs. PPV23-alone (base-case ICER of C$44,324/QALY); **B**. PCV20-alone vs. PPV23-alone (base-case ICER of C$46,961/QALY); **C**. PCV20-alone vs. PCV15-alone (base-case ICER of C$70,751/QALY). The x-axis depicts the range of the ICER values (C$/QALY). CAP= Community-acquired pneumonia; ICER = Incremental cost-effectiveness ratio; IPD=Invasive pneumococcal disease; NBPP= Nonbacteremic pneumococcal pneumonia; PCV15 = 15-valent pneumococcal conjugate vaccine; PCV20 = 20-valent pneumococcal conjugate vaccine; PPV23 = 23-valent pneumococcal polysaccharide vaccine; QALY = Quality-adjusted life year; ST = Serotype 3.

## DISCUSSION

We evaluated the cost-effectiveness of high-valency PCV (PCV15 and PCV20) vaccines for adults aged 65 years and older from a healthcare payer perspective in Ontario, Canada. The findings showed that vaccinating older adults with PCV15 or PCV20 vaccines reduced the incidence of pneumococcal disease and mortality, resulting in QALY gains, albeit at a higher cost compared to the current routine pneumococcal vaccination program using PPV23.

Our results are consistent with previous Canadian study by the Public Health Agency of Canada, which showed that vaccinating older adults with PCV15 or PCV20 alone is likely to be cost-effective compared to using PPV23-only, with ICERs below C$50,000/QALY [31]. When comparing all strategies sequentially at a $50,000/QALY threshold, both studies indicated that PCV15/20 in series with PPV23 may not be costs effective options; however, contrary to our findings, the Public Health Agency of Canada study reported that PCV15 alone was dominated (more costly and less effective) compared to PCV20 [31]. ICER disparities between these studies could be attributed to variations in key parameter estimates such as vaccine price, pneumococcal disease burden, vaccine effectiveness and waning. Our sensitivity analyses explored the impact of these assumptions, providing valuable insights for decision-making under different scenarios.

Our findings are also comparable to those from US studies [41] and a systematic review conducted by the Public Health Agency of Canada [31]. Three US studies from the healthcare perspective found that the cost-effectiveness of PCV20-alone compared to PPV23-alone ranged from cost-saving (lower cost and improved health outcomes) to an ICER of US$39,000/QALY gained [41]. Adding PPV23 to PCV15 or PCV20 resulted in ICERs ranging from cost-saving to US$282,000/QALY gained when compared to PPV23. Notably, the Centers for Disease Control and Prevention (CDC) model found that PCV20-alone or PCV15 in series with PPV23 was cost-savings in all scenarios considered [41]. Another US study reported a higher ICER for PCV20-alone (US$172,491/QALY) compared to PPV23, while PCV15-based strategies were dominated by PCV20 [42]. This study echoed our findings that PCV20 in series with PPV23 was not cost-effective when compared to PCV20-alone, with an ICER exceeding US$3 million/QALY [42]. The systematic review reported that ICERs for PCV20 alone compared to PPV23 ranged from cost-saving to $187,761/QALY, while ICERs for PCV15 alone versus PPV23 was $250,434/QALY [31]. Similar to our study, combined use of PCV15 or PCV20 with PPV23 was unlikely to be cost-effective, with ICERs surpassing commonly used thresholds [31]. The substantial difference in ICERs between the studies can be explained by differences in key model input estimates, highlighting the importance of context-specific data to make informed decisions.

Our study found that PCV15/20 vaccines (alone or in series with PPV23) may become less economically attractive for older adults in the long-term when considering their potential use in childhood vaccination programs and associated indirect (herd immunity) effects, which is consistent with results of the Public Health Agency of Canada modelling study and the systematic review [31]. Similarly, other studies generally reported higher ICERs when indirect effects were considered [19, 41, 42]. This demonstrates the need to consider comprehensive pneumococcal vaccination program options across age and risk groups when developing recommendations.

Because real-world data were unavailable at the time of the analysis, there was significant uncertainty on some parameter values for PCV15 and PCV20. The base-case results were most sensitive to vaccine price, vaccine effectiveness, waning of vaccine effectiveness, and vaccine-preventable burden of pneumonia, which were also reported as influential parameters in other studies [31]. The US study demonstrated that lower vaccine prices or increased vaccine effectiveness resulted in reducing the ICER to <$100,000/QALY gained [43]. Overall, our study indicated that PCV15/20 serotype coverage for older adults is still suboptimal, resulting in small incremental health benefits compared to current routine vaccination with PPV23.

Our study has some limitations. As stated, there was uncertainty about some PCV15/20 parameter values such as vaccine cost and effectiveness; however, our broad sensitivity and scenario analyses can inform decision-making as more evidence becomes available. We also assumed that PPV23 has no effect in preventing NBP cases as studies yielded inconclusive results which is a conservative approach; however, this assumption may bias the analyses in favour of strategies that only use PCV vaccines without PPV23. Should PPV23 be effective against NBP, the new PCV vaccinations would become less economically attractive. We were unable to account for some potential benefits such as avoiding increasing frailty post *S*.*pneumoniae* infection, which could result in lower quality of life and higher healthcare cost, including potential admission to long-term care. Due to data limitations, we included the indirect effect of pediatric pneumococcal vaccination only in the sensitivity analysis, estimating it as a percentage reduction in pneumococcal disease. However, this approach may oversimplify the overall impact of the vaccines, especially since serotype replacements were not modeled. Further, our study was conducted from a healthcare payer perspective, not from a societal perspective, and as a result, we excluded out-of-pocket expenditures, caregiver burden, and productivity losses. We did not model the costs and QALYs related to vaccine adverse events, though including them is unlikely to substantially change the overall estimates, given that for pneumococcal vaccines these events are expected to be rare, mild, and generally not necessitate healthcare intervention. Further, the vaccines are considered to have comparable safety profile.

Despite these shortcomings, we performed a thorough economic analysis using real-world age-specific disease incidence and cost data from the province’s population-based datasets. Our findings could assist Ontario in making informed recommendations on the use of the new pneumococcal vaccines, considering the cost-effectiveness results and the broader impact on the entire program, e.g., the potential for herd immunity effects. Our findings can also inform deliberations in jurisdictions with similar pneumococcal disease epidemiology, existing vaccination programs and healthcare systems.

## CONCLUSIONS

Our study indicates that vaccination of adults 65 years and older with PCV15 or PCV20 alone is cost-effective compared to the current vaccination program with PPV23 at a cost-effectiveness threshold of C$50,000/QALY. However, use of PCV15 or PCV20 in series with PPV23 is unlikely to be cost-effective. Cost-effectiveness may be impacted in the future when PCV15 or PCV20 use in childhood vaccination programs, is expected to result in indirect protection of adults. The most influential parameters in the sensitivity analyses were vaccine cost, vaccine effectiveness, vaccine effectiveness waning, and proportion of community-acquired pneumonia caused by *S. pneumoniae*.

## Data Availability

All data produced in the present work are contained in the manuscript.

## Funding

This study was funded by a University of Toronto Connaught Global Challenge Award 2019–2020. GBG also received a postdoctoral fellowship award from the Center for Vaccine Preventable Diseases at the Dalla Lana School of Public Health. The Center for Vaccine Preventable Diseases receives funding from government, philanthropic, not-for-profit and private sector organizations, including vaccine manufacturers. This includes a donation from Sanofi to Center for Vaccine Preventable Diseases. A donation from Merck to the Center for Vaccine Preventable Diseases, at the Dalla Lana School of Public Health, supported the salary of SAF from April 2020 to February 2023. SAF does not receive funding directly from Merck or any personal payment or direct funds from vaccine manufacturers to support her research. The funder had no role in study design, implementation, data analysis and interpretation. All authors attest they meet the ICMJE criteria for authorship. This research was also supported, in part, by a Canada Research Chair in Economics of Infectious Diseases held by Beate Sander (CRC-2022-00362).

## Author contributions

GBG performed literature search, developed the model, conducted the economic analysis and drafted the manuscript. RO contributed to data curation and economic analysis. SM contributed to economic analysis. BS supervised and guided data curation and the economic analysis. GBG, NSC, SAF and BS solicited the funding. All authors provided methodological input, interpreted the results, critically reviewed the manuscript, and have approved the final version for publication.

## Declaration of Competing Interest

The Centre for Vaccine Preventable Diseases is operated by the Dalla Lana School of Public Health, which receives funding from government, philanthropic, not for profit and private sector organization, including vaccine manufacturers. This includes a donation from Merck to the institution that supports the salary of SAF. SAF does not receive funding directly from Merck or any personal payment or direct funds from vaccine manufacturers to support her research. Other authors declare no conflicts of interest.

## Notes

### Competing Interest Statement

The authors have declared no competing interest.

